# Landscape of Tandem Repeat Variations in Multi-ethnic Asian Populations

**DOI:** 10.64898/2026.08.03.26359643

**Authors:** Qi Jia, Max Lam, Ling Wang, Feifan Zhao, Huayang Tang, Prasad Sarashetti, Zhe Li, Eleanor Wong, SG10K_Health Consortium, Patrick Tan, Xueling Sim, Joanne Ngeow, Jimmy Lee, Ching-Yu Cheng, Miao Ling Chee, Weng Khong Lim, Calvin Woon Loong Chin, Neerja Karnani, Yap Seng Chong, Wei Cheng Sim, Chia Wei Lim, Nicolas Bertin, Jianjun Liu

## Abstract

Tandem repeats (TRs) are implicated in over 70 Mendelian disorders and likely contribute to the “missing heritability” of complex traits and diseases, yet TR variations in Asian populations remain poorly characterized. Here, we constructed an Asian-specific SG10K-TR catalog by leveraging the SG10K_Health Dataset, comprising 916,274 autosomal TR loci genotyped in 9,490 individuals of Chinese (5,528), Malay (1,824), Indian (2,108), and other ancestries (30). Using a novel integrative measure for both repeat length and frequency variations, TRDDS, we found that population-level TR variations are selectively constrained in coding and promoter regions, whereas the enrichment of TRs with high population diversity was observed in regulatory sites with low chromatin accessibility and pathways related to neuronal functions. We also identified candidate TRs under selection that predominantly targets neuronal and synaptic architecture. Analysis of linkage disequilibrium (LD) patterns revealed that TRs are often poorly tagged by small variants, although we identified 123 candidate functional TRs that may underlie association signals previously attributed to nearby noncoding SNPs. Finally, TR-based GWAS of six anthropometric and lipid traits identified ten loci with genome-wide significant associations, including two novel loci for BMI (*LINC02817*) and height (*UNC45B*), and a TR variant as causal candidate for a known GWAS locus at *HMGCR* for LDL. Together, this study establishes a critical Asian-specific TR resource and highlights the fundamental role of TR diversity in driving evolutionary neuroplasticity and shaping the genetic architecture of complex traits.

## Introduction

Tandem repeats (TRs) are repetitive DNA elements that comprise 3%-5% of the human genome, including short tandem repeats (STRs) and variable number tandem repeats (VNTRs)^1^. Their repetitive architecture predisposes TRs to strand slippage during DNA replication, leading to a high mutation rate estimated at 4.74 × 10^-6^ per locus per haplotype per generation^2^, resulting in high degrees of polymorphism. To date, more than 70 Mendelian disorders have been attributed to pathogenic TR variation^3^. Beyond monogenic diseases, TRs have also been increasingly implicated in the genetic susceptibility of complex traits and diseases^4,5^. Emerging TR-based genome-wide association studies (GWAS) across traits such as anthropometric measurements^6^, blood serum biomarkers^7^, and various neurological disorders^8,9^ further suggest that TR variation may account for part of the “missing heritability” where SNP-based GWAS fails to explain^10^.

A robust and population-diverse TR catalog is essential for advancing TR research. Several large-scale datasets have contributed to TR characterization, such as the 1000 Genomes Project (1KGP)^11,12^ and the NyuWa dataset^13^, the latter of which represents a large but exclusively Chinese cohort. The largest resource to date, the TR-Atlas, includes more than 300,000 individuals^14^; but still predominantly reflects European, African, and Hispanic ancestry, with only limited representation of East and South Asian populations. Consequently, TR variation across East, Southeast, and South Asian ancestries remains insufficiently characterized.

Singapore’s multi-ethnic population, which comprises Chinese, Malay, Indian, and other groups, provides a valuable opportunity for addressing this gap. The Singapore National Precision Medicine (NPM) initiative is a three-phase national program that integrates genomic, lifestyle, health, and environmental data to advance precision medicine^15^. NPM Phase I generated the SG10K_Health dataset, substantially expanding our understanding of genomic diversity in Asian populations^15^. For example, structural variation (SV) analysis using the SG10K_Health Dataset identified 47,770 novel SVs^16^, which represent a key class of complex variations alongside TRs.

In this study, we leveraged the SG10K_Health cohort to characterize TR variation in Asian populations and to conduct TR-based GWAS. These analyses provide deeper insights into TR diversity in Asian populations and revealed contributions of TRs to several complex traits. Our study collectively establishes an important resource for future genetic and clinical research.

## Results

### The SG10K-TR catalog

We conducted TR genotyping of 9,490 individuals by using whole-genome sequencing (WGS) data generated through the SG10K_Health study^15^. This cohort includes three major ethnic groups in Singapore: 5,528 Chinese (58.3%), 1,824 Malay (19.2%), and 2,108 Indian (22.2%), along with 30 individuals of other ancestries (0.3%). The WGS data were produced using different sequencing protocols regarding library preparation and sequencing depths. Specifically, 6,045 individuals were sequenced using PCR-based library preparation at a target depth of 15x (15x_PCR-based), 1,922 using PCR-based at 30x depth (30x_PCR-based), and 1,523 using PCR-free preparation at 15x depth (15x_PCR-free). TR genotyping for the SG10K_Health dataset largely followed the EnsembleTR framework developed by Ziaei Jam *et al.*^11^ GRCh38-aligned CRAM files were processed with three TR callers, including Expansion Hunter (EH)^17^, GangSTR^18^, and HipSTR^19^, to profile autosomal TRs (motif length ≥ 2bp). Genotyping results from individual callers were harmonized using EnsembleTR^11^, with two layers of quality control filtering^20^ applied before and after harmonization (Fig. 1a, Methods).

**Figure 1:**
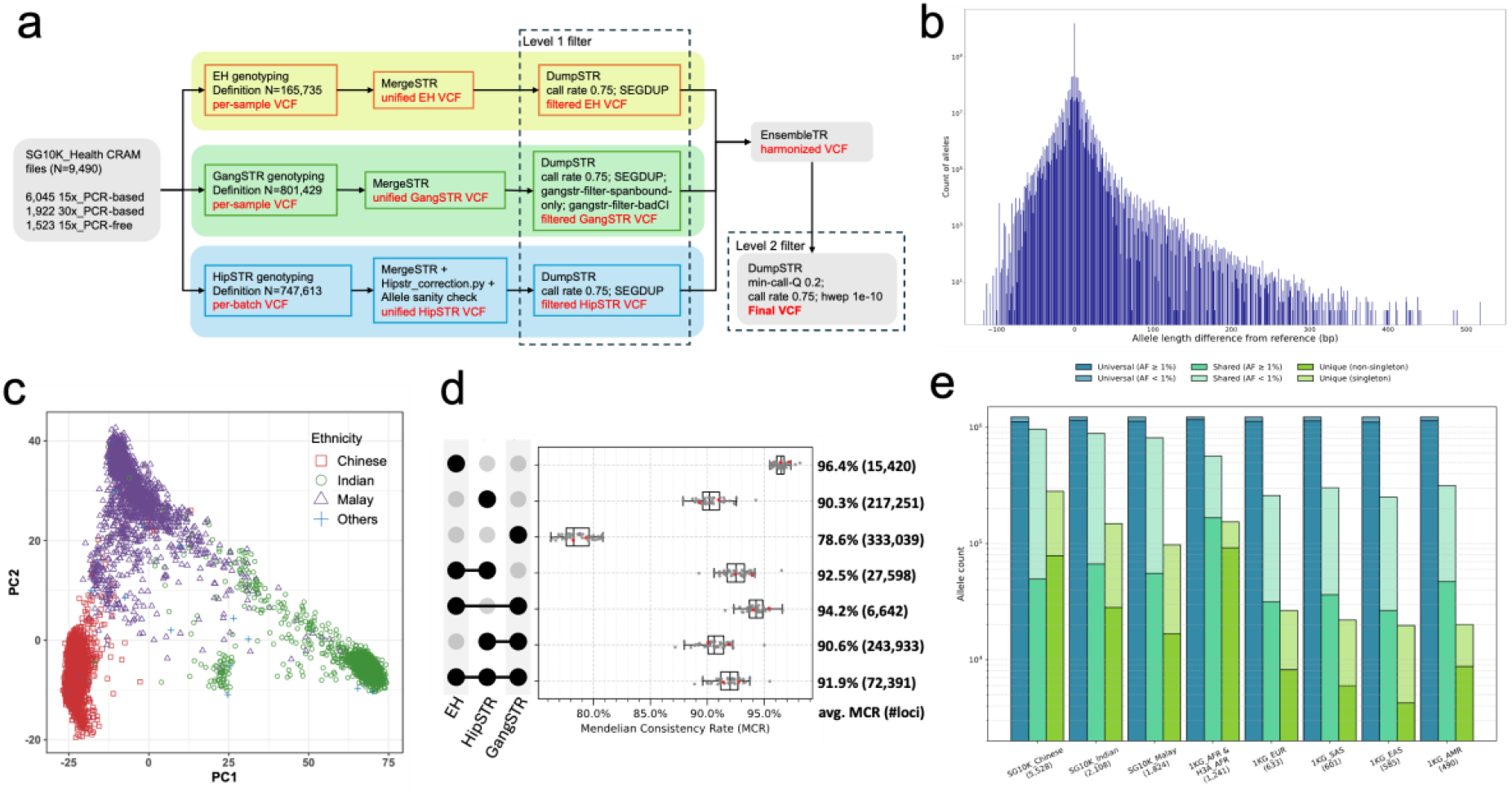
Overview of the SG10K-TR catalog. **a** Workflow of generating SG10K-TR catalog. **b** Distribution of TR allele lengths relative to the reference in the SG10K-TR catalog. **c** Principal component analysis (PCA) based on TR genotypes reveals the population structure of SG10K individuals. **d** Number of TR loci supported by individual TR callers and their combinations after harmonization by EnsembleTR, together with the corresponding Mendelian consistency ratios (MCR) based on 45 trios. Red dots indicate trios containing samples sequenced by different protocols. **e** Allele sharing between the 1KG_H3A-TR catalog and SG10K-TR catalog.

The resulting SG10K-TR catalog comprises 916,274 autosomal TR loci, with repeat unit length ranging from 2 to 24 bp. Among these, 660,157 loci exhibited TR length variation among individuals and were classified as polymorphic TRs (pTRs); the remaining loci appear monomorphic in the current cohort, although additional alleles may emerge with increased sample size or in other populations. In addition, 189,375 loci harboured more than one common allele (AF ≥ 0.01). TR variations are primarily driven by insertions or deletions of whole or partial repeat units, manifesting as indels relative to the defined TR regions on the reference genome. Alleles in the SG10K-TR catalog exhibited length differences from -116 bp to +518 bp, or -46 to +221 repeat units relative to reference alleles based on GRCh38 (Fig. 1b). The distribution of TR allele lengths exhibits a two-tiered asymmetry. Firstly, large insertions (>100 bp) were more frequently observed than large deletions of comparable size. This likely reflects a natural constraint of reference- and definition-based TR genotyping. Deletions have a low bound because the deletion can only be as large as the one that completely deplete of the defined TR sequence. In contrast, repeat insertions are not subject to an equivalent upper bound. Secondly, the number of repeat deletion events were higher than the ones of the insertions, with an average of 76,503 insertion events and 82,715 deletion events captured per individual. Further stratification of indel events by reference allele length revealed that number of deletion events surpass insertion events particularly when the reference allele length exceeds 40 bp (Fig. S1). Moreover, a gradual decline of mean allele length as the length of reference allele is increasing^20^, with a notable drop for loci with reference alleles exceeding 120 bp (Fig. S2). These suggest a “definition-dependent bias” of TR genotyping likely compounded by short-read limitations, leading to an underrepresentation of insertions in long-spanning TR loci.

Principal component analysis (PCA) of TR alleles confirmed expected population structure along PC1 and PC2, clearly separating Chinese, Malay, and Indian ethnic groups (Fig.1c). However, higher-order components, including PC3 and PC5, revealed stratification by sequencing protocol groups, indicating the presence of effects driven by sequencing depths and PCR steps (Fig. S3). To mitigate potential confounding, downstream analyses were performed using the pooled dataset, but key findings were further examined within each sequencing protocol group (Supplementary Methods).

To assess genotyping accuracy, we evaluated Mendelian consistency rate (MCR) using 45 parent-offspring trios from the cohort, achieving an overall MCR of 91.2% (Fig. 1d). Similar to the previous report^11^, TRs called exclusively by GangSTR showed reduced accuracy, whereas integrating results from multiple callers generally improved MCR (Fig. S4). Notably, two trios whose samples were sequenced with different protocols exhibited high MCRs (92.1% and 90.7%), indicating that the catalog is robust to technical differences across sequencing protocols.

We compared the SG10K-TR catalog with the only open-source, population-scale TR callset based on samples from 1000 Genomes Project and H3Africa (N = 3,550)^11^, hereafter referred to as the 1KG_H3A TR catalog. Among the 835,072 TRs exactly shared between the two catalogs, the SG10K-TR catalog substantially increased allelic diversity across loci (Fig. 1e). Specifically, the three major SG10K ethnic groups show a higher proportion of unique alleles (SG10K Chinese: 11.4%, SG10K Malay: 4.6%, SG10K Indian: 6.6%) compared to the 5 superpopulations in the 1KG-H3A TR catalog, with the exception of the African group (7.9%). In total, 526,891 unique alleles were identified in the SG10K-TR catalog, of which 123,482 are non-singletons (Supplementary Data 1).

### TR diversity among three ethnic groups

To assess TR diversity among Asian populations, we employed two complementary metrics. Rst, developed by Slatkin^21^, is a widely used measure of population subdivision for microsatellite alleles and is conceptually analogous to Wright’s fixation index (Fst) for SNPs. In contrast, the Tandem Repeat Disparity Score (TRDS), introduced by Cui *et al.*^14^, quantifies differences in TR allele distributions between two groups using the 2-Wasserstein distance. Both metrics were computed for each pTR across three major ethnic group pairs Chinese-Malay (CM), Malay-Indian (MI), and Indian-Chinese (IC), as well as for the three major ethnic groups combined (overall; see Methods).

Although Rst(overall) and TRDS(overall) exhibited a strong global association (Pearson’s r = 0.81 on log-transformed values), the correlation was reduced when considering individual population pairs, such as Rst(CM) and TRDS(CM) (r = 0.66; Fig. S5). Despite this overall correlation, concordance among top-ranked loci was limited, with only 24% of loci overlapping between the top 1% ranked by Rst(overall) and TRDS(overall), indicating that the two metrics are likely to prioritize distinct extreme loci. Inspection of the allele distribution revealed a general tendency whereby Rst is driven primarily by allele frequency differences and repeat-length variation, whereas TRDS is more sensitive to distributional shifts, including rare but large repeat expansions or contractions. Allele distributions of the top ten loci are shown in Fig. S6 for illustration. Since Rst and TRDS capture partial yet complementary features of TR variation, we therefore defined a novel composite metric, the Tandem Repeat Differentiation Disparity Score (TRDDS), as the product of Rst and TRDS. TRDDS showed strong associations with both component metrics across overall and population pairs analyses, while improving balance between allele frequency differentiation- and distribution-driven signals (Fig. S5, Fig. S7). TRDDS thus provides a more holistic measure for quantifying TR variation diversity across populations.

To evaluate the functional impact of TR diversity across populations, we examined the relationship between the TRDDS(overall) and gene annotation and candidate cis-regulatory region (cCRE) annotations^22,23^ (Fig. 2a,b; Fig. S8). In general, TRs located in coding sequences (CDS), 5′ untranslated regions (5′UTRs), and promoter-like sequences (PLS) tended to be more conserved, consistent with stronger selective constraints due to their direct roles in gene expression. In contrast, population-level TR diversity is elevated in transcription factor binding sites exhibiting low chromatin accessibility (TF (low CA)). This pattern suggests that TR diversity preferentially accumulates in regulatory elements with weaker or context-dependent activity rather than in constitutively active regulatory regions, since chromatin accessibility is dynamically modulated by external stimuli and developmental cues^24^.

**Figure 2:**
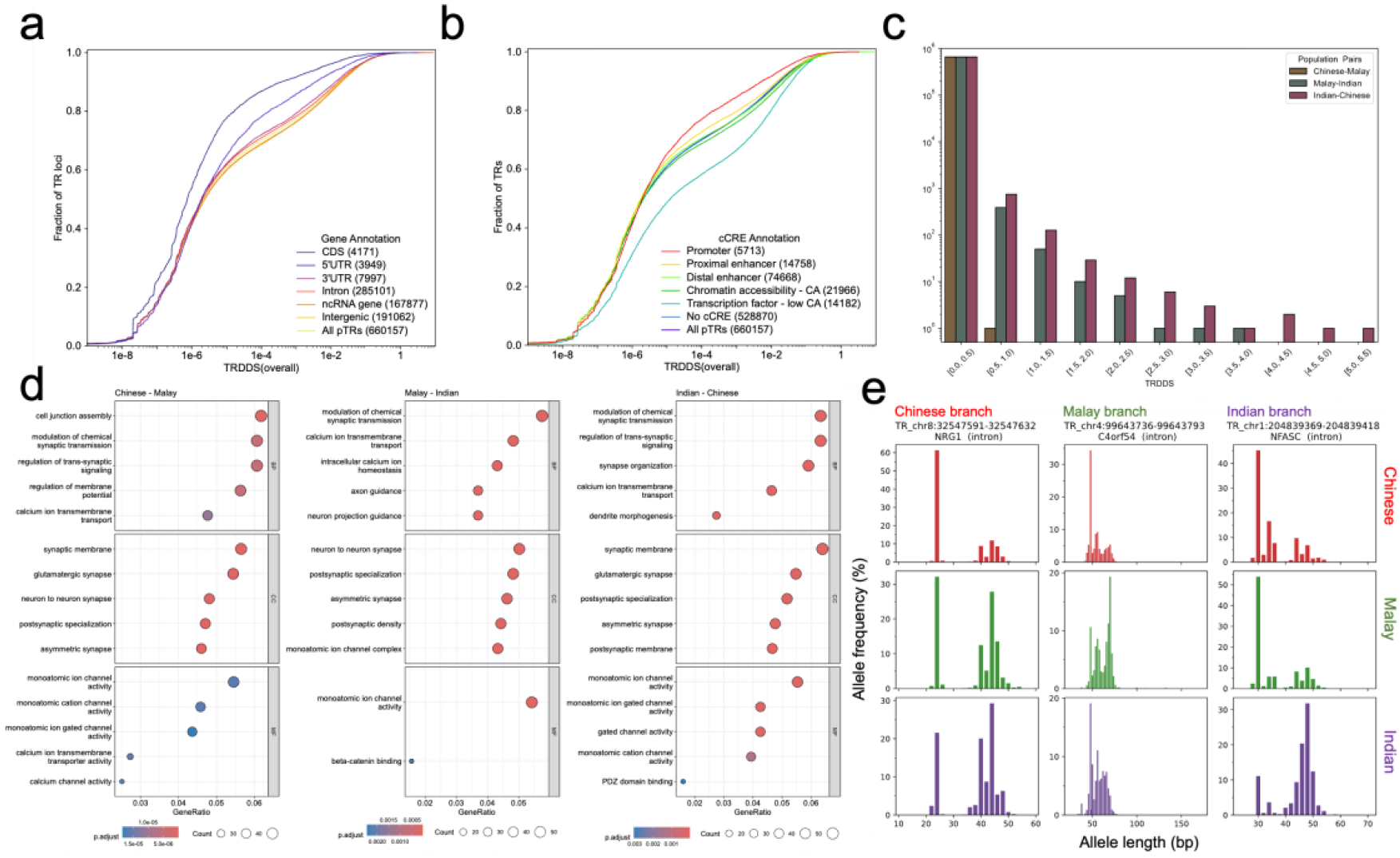
TR diversity among three ethnic groups. **a,b** Cumulative distribution of TRDDS(overall), stratified by gene region annotation (a) and cCRE annotation (b). **c** Distribution of TRDDS across three population pairs - CM, MI and IC. **d** Gene Ontology (GO) enrichment of genes overlapping the top 0.5% of TRDDS loci for three population pairs. **e** Allele length distributions of TR loci under selection in Chinese, Malay, and Indian branch.

Comparisons of TRDDS among population pairs revealed differences in distribution. In particular, IC and MI exhibited a broader TRDDS distribution extending to higher TRDDS bins, whereas CM was largely confined to lower TRDDS ranges (Fig. 2c; Fig. S9). this pattern is consistent with previously reported divergence times, where SG Indians diverged from SG Chinese (∼45.9 kya) and SG Malays (∼43.9 kya) earlier than the split between SG Chinese and SG Malays (∼24.8 kya)^25^.

We then conducted GO enrichment using genes whose sequences overlapped with the significant (top 0.5%) TRDDS loci for CM, MI and IC. We identified 89, 57 and 68 enriched pathways for CM, MI and IC, respectively (Fig. 2d; Supplementary Data 2). Notably, despite exhibiting the lowest overall genetic divergence, CM revealed the strongest functional enrichment in both number of enriched pathways and statistical significance. This pattern suggests that the relatively limited TR differentiation between Chinese and Malay populations is concentrated in specific biological process. In contrast, the broader functional dispersion in IC and MI enrichments may result from signal dilution due to deeper evolutionary divergence, where both selective and neutral processes may contribute to TR diversity.

In total, there are 30 enriched pathways were shared across all three pairs, and 29 pathways share by two pairs. Dominant themes among shared pathways are related to synaptic and neuronal functions, including synaptic transmission, dendrite development, calcium transport and specialized membrane architecture. These shared enrichments indicate that population-level TR diversity recurrently impacts basic neuronal signalling and ion channel biology. To provide locus-level examples underlying the enrichment of these calcium transport pathways, we identified 12 TR loci across seven calcium channel complex genes that are significant in at least one population pair (Fig. S10). Pair-specific pathways revealed distinct functional nuances. CM-unique enrichments were characterized by broader structural and developmental processes (e.g., cell-substrate adhesion, embryonic morphogenesis), whereas deeper IC and MI divergences drilled into highly specialized neurological domains (e.g., presynaptic active zones, telencephalon development). This distribution suggests a temporal dynamic in TR evolution that, while TRs may initially facilitate a broad spectrum of morphological and basic neuronal adaptations, the predominant, long-term selective pressure driving TR divergence is overwhelmingly dedicated to highly specialized neurodevelopmental and synaptic fine-tuning.

Having established the systemic, pathway-level impact of population TR diversity, we next sought to identify candidate loci under selection. To infer distinct evolutionary trajectories, we calculated a TR-based Population Branch Statistic (TR-PBS) derived from pairwise TRDDS values. By applying a threshold isolating the top 0.1% maximum TR-PBS values across the three populations and requiring a fold change ≥ 2 to filter out shared ancestral events, we defined a set of 508 candidate loci under selection (Fig. 2e; Supplementary Data 3). The vast majority (N = 506) of these highly divergent loci fall on the Indian branch, though we also identified one TR locus specific to the Chinese branch and one to the Malay branch. To note that because we did not include an outgroup, our TR-PBS calculation relies on an unrooted tree topology. Therefore, loci assigned to the Indian branch may represent positive selection specific to the South Asian lineage, or selection that occurred in the shared ancestral population of the Chinese and Malays prior to their split^25^.

The candidate locus on the Chinese branch (TR_chr8:32547591-32547632) is located within an intron of *NRG1* gene, which encodes a critical ligand for the ERBB signalling pathway that plays a central role in diverse neuronal activities^26,27^. The candidate on the Malay branch (TR_chr4:99643736-99643793) resides within an intron of *C4orf54*. While the endogenous function of *C4orf54* remains understudied^28^, this TR overlaps with a candidate distal enhancer that interacts with the core lipid metabolism gene *MTTP*^29^ in left ventricle tissue (SCREEN^22,23^ accession: EH38E3596757). Supporting this physical interaction, we also found that SNPs in moderate linkage disequilibrium (LD) with this TR in Chinese and Malay primarily tag established GWAS signals for body anthropometrics^30^ and MHPG level pertinent to the central nervous system^31^ (Supplementary Data 4). Together, these indicate that this TR under positive selection along the Malay lineage likely reflects the pleiotropic regulatory roles in both systemic metabolism and neurochemistry. Looking at the deep divergence separating the Indian lineage, the top-ranked loci are located in genes highly related to neuronal functions, such as *NFASC*^27^, *RASGRF2*^32^, and *KATNA1*^33^. Moreover, further GO enrichment of the genes overlapping candidate loci revealed pathways exclusively dedicated to core neurobiology and synaptic architecture, including the main axon, postsynaptic specialization, and GABA-ergic synapses (Supplementary Data 5). Interestingly, several genes harbouring our TR candidates under selection, including *OPCML*, *TENM2*, and *CDH13*, have been previously implicated in autism spectrum disorder (ASD) via TR expansions, though at distinct loci^34^, which underscores the exquisite sensitivity of these neuronal networks to TR-mediated regulation.

### Linkage Disequilibrium among TRs

Linkage disequilibrium (LD) patterns involving TRs and small variants (SNPs and short indels) were examined across the autosomes. TR genotypes were encoded into relative diploid TR dosage for LD calculation to better reflect the nature of TR variations in length (Method). Overall, compared to LD (small variant, small variant), LD(TR, small variants) was approximately half the strength, while LD (TR, TR) was about a third (Fig. 3a; Fig. S11). This pattern is consistent with the previous observations on chromosome X^12^. Across the three ethnic groups studied, Indian individuals exhibited the fastest decay of LD (TR, small variant), followed by Malay and Chinese, suggesting higher TR diversity in the Indian population (Fig. 3b; Fig. S12).

**Figure 3:**
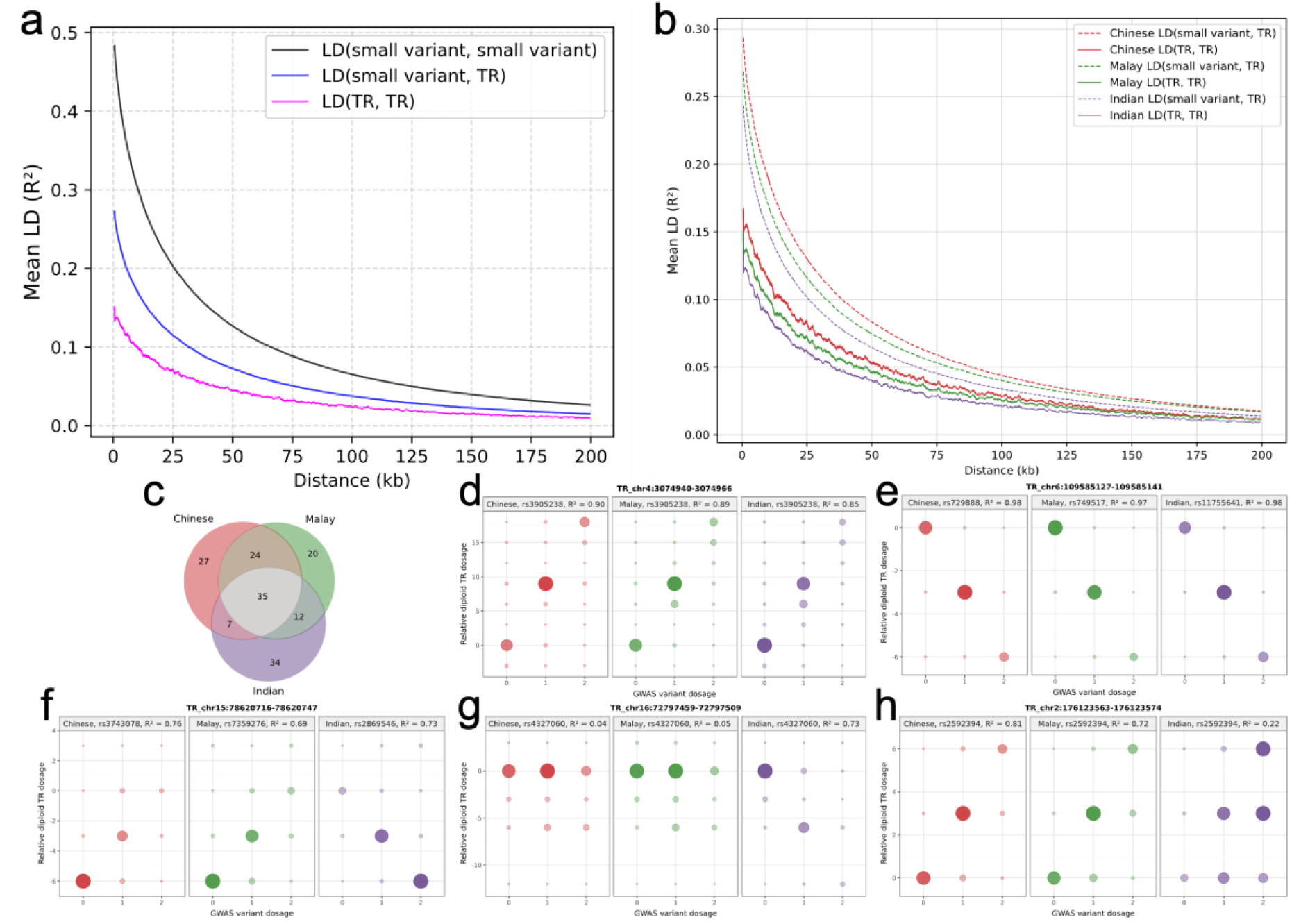
TR Linkage Disequilibrium (LD) patterns. **a** Overall unphased LD decay pattern among small variant and TR. **b** Unphased LD(small variant, TR) and LD(TR, TR) decay patterns across Chinese, Malay and Indian populations. **c** Number of LD pairs involving candidate functional TRs and GWAS variants shared among the three populations. **d-h** Dot plots of relative diploid TR dosage against GWAS variant dosage for three populations at five representative TR loci: **d** TR_chr4:3074940-3074966, **e** TR_chr6:109585127-109585141, **f** TR_chr15:78620716-78620747, **g** TR_chr16:72797459-72797509, and **h** TR_chr2:176123563-176123574. Relative diploid TR dosage categories with population frequency < 0.01 were excluded from visualization to align with the filter applied for LD r^2^ calculation.

Having characterized the overall LD patterns of TRs, we next explored their contributions to GWAS findings, particularly in terms of their potential roles as functional causal variants of previous GWAS loci. Because GWAS signals often implicate markers that are in high LD with causal variant, we examined TRs in high LD (r² ≥ 0.6) with variants reported in the GWAS Catalog to assess their potential contribution to trait-associated signals. This yielded 219,199 LD pairs, with 165,175 LD pairs for Chinese, 148,894 LD pairs for Malay, and 135,809 LD pairs for Indian. These high LD pairs span 52,391 unique TRs and 96,605 unique GWAS variants. To further prioritize candidate functional TR loci, we focused on LD pairs in which the TR is located within CDS, while the linked GWAS variant does not directly impact the CDS. Because individual CDS-located TRs were often in high LD with multiple nearby GWAS variants, we retained only the GWAS variant with the highest r² per TR locus to obtain a single representative TR - GWAS variant LD pair, thereby reducing redundancy. Using this approach, we identified 93, 91, and 88 LD pairs involving candidate functional TRs in Chinese, Malay, and Indian populations, respectively (Fig. 3c; Supplementary Data 6), comprising 123 unique candidate functional TRs and 154 linked GWAS variants.

Among these candidate functional TRs, 35 were shared across all three populations with the same tagged GWAS variants, and 31 TRs were shared across three populations but associated with different tagged GWAS variants. Applying an additional stringent filtering criterion to identify population-specific signals (Methods), we further identified three Indian-specific candidate functional TRs and one candidate functional TR shared between Chinese and Malay populations (Fig. 3d-h; Fig. S13).

TR_chr4:3074940-3074966, which encodes the polyproline (poly P) tract of the huntingtin protein (*HTT* gene) is in strong LD for all three populations with rs3905238 (Fig. 3d), which is associated with depressive disorders^35^. This poly P tract lies adjacent to the well-known polyglutamine (poly Q) tract implicated in Huntington’s disease^36^. We identified a TR_chr6:109585127-109585141 which encodes for poly glutamic acid (poly E) tract in the *AK9* gene that shows high LD with three SNPs for each population (Fig. 3e), all of which reside within an acute myeloid leukemia (AML) risk haplotype reported before^37^. While the original GWAS work identified the haplotype as a whole, our finding that the same TR consistently links to different components of this haplotype across populations suggests it is a core functional element underlying this haplotype’s association with AML. TR_chr15:78620716-78620747, which encodes for a polyleucine (poly L) tract in *CHRNA3* gene, is also in high LD with different GWAS SNPs for each population (Chinese: rs3743078; Malay: rs7359276; Indian: rs2869546; Fig. 3f), but the GWAS SNPs are associated with different traits, where rs3743078 is primarily associated with schizophrenia^38,39^, rs7359276 is associated with response to bronchodilator^40^ and rs2869546 is associated with velopharyngeal dysfunction^41^. This indicates potential pleiotropic effects of this TR locus. For population specific LD pairs, TR_chr16:72797459-72797509 maps to a poly Q tract in *ZFHX3* gene, and it is in high LD with rs4327060 in Indian populations that are associated with vitamin D amount^42^, but with minimal LD with Chinse and Malay (Fig. 3g). Inversely, TR_chr2:176123563-176123574 encodes for poly Q tract in *HOXD9* gene, and it is in high LD with rs2592394 in Chinese and Malay only (Fig. 3h). rs2592394 is associated with several traits such as magnesium measurement^43^ and peripheral arterial disease^44^.

### TR GWAS

Finally, we performed the TR-based GWAS on six anthropometric and lipid traits in the SG10K_Health dataset, including body mass index (BMI), Height, high density lipoprotein level (HDL), low density lipoprotein (LDL), total cholesterol level (TC) and triglyceride level (TG). After sample QC (Methods), 6,458 individuals were retained, with the exact number of samples per trait depending on phenotype availability (Supplementary Data 7). For TR GWAS, TR genotypes were represented as the relative diploid TR dosage, defined as the total allele-length difference from the reference length (Methods). Association testing was performed using normalized trait values and the TR genotypes in a linear regression model with adjustments for sex, age and PC1-PC5. Across the six anthropometric and lipid traits, an average of 195,942 TR loci (194,703-196,543) were tested per trait after QC and frequency filtering. The genomic inflation factors were small (λ = 1.009-1.033; Supplementary Data 7), indicating that test statistics were well-calibrated without inflation. A genome-wide significance threshold of P < 2.5 × 10^-7^ was defined for TR GWAS based on ∼200k tests per trait. In total, ten genome-wide significant associations within nine TR loci across five traits were identified, with no genome-wide significant associations observed for TC (Supplementary Data 8).

Ten genome-wide significant TR associations were cross-referenced with the GWAS Catalog for the corresponding traits to identify nearby known associations. Nine of the ten associations were located within ±100 kb of previously reported GWAS signals. The BMI-associated TR at chr1:221335333-221335386 P = 2.49 × 10^-7^, β = 0.017; Fig. S14) is a novel locus that is not overlapping with any reported GWAS associations. This TR variant is located within a long non-coding RNA *LINC02817*. In addition, this TR also overlaps a candidate proximal enhancer (SCREEN^18,33^ accession: EH38E1424686) with regulatory activity observed predominantly in brain tissues, with additional supporting evidence in liver and muscle tissues. Inspection of the reference sequence shows that this locus primarily contains two adjacent repeat segments-(GT)n and (AG)n, producing a complex repeat architecture. When these repeat segments are represented as individual variants, their association signals are weaker than the TR-based test (Fig. S15), suggesting that TR-based genotype are able to capture the combined effects of multiple variants within the locus.

Among the nine TR associations with nearby reported GWAS signals, eight were in moderate to high LD with known variants (r² ≥ 0.4). In contrast, TR_chr17:35177802-35177914, located in the intron of *UNC45B* and associated with height (P = 1.65 × 10^-7^, β = 0.041; Fig. S16), appears to represent a novel signal that is independent of previously reported associations. The only nearby reported variant, rs12939024^25^ in *SLFN5*, showed only marginal association in the current dataset (P = 5.89 × 10^-2^, β = - 0.107) and was in minimal LD with the TR (r² = 0.0011; Fig. S17). This repeat is dominated by the motif CTTTT, interspersed with imperfect C(T)n units, producing a locally complex repetitive sequence context. The observed TR variation can be decomposed into two discrete indels: a 5 bp deletion (rs201627371, P = 1.64 × 10^-6^, β = -0.18) and a 1 bp intra-repeat deletion (rs80168474, P = 5.52 × 10^-1^, β = -9.6 × 10^-3^). Individually, these indels show weaker or non-significant associations with height, but the TR length-based genotypes aggregate these sequence changes into a single variable, producing a stronger association signal than the individual variants being tested separately. The repeat region overlaps a predicted distal enhancer (SCREEN^22,23^ accession: EH38E3219212) that interacts with *SLFN5* and *SLFN12L*, suggesting a potential regulatory function.

In addition to the two novel signals, a TR within the intron of *HMGCR*, TR_chr5:75352250-75352286, was identified as a stronger marker for LDL levels (P = 1.11 × 10^-9^, β = 0.0078) with more significant association evidence than other nearby variants (Fig. 4a-c). *HMGCR* gene encodes a rate-limiting HMG-CoA reductase in cholesterol biosynthesis and has been widely implicated in lipid metabolism^45,46^. Multiple known GWAS variants are located in this region, including rs10056811^47^ (P = 3.45 × 10^-9^, β = 0.11) and rs6882842^48^ (P = 1.44 × 10^-7^, β = -0.094), which are in moderate LD with the TR but show weaker associations in the current dataset (Fig. 4e). The TR consists of a repeated ATT motif and is highly polymorphic, with 18 alleles spanning a range of repeat lengths (Fig. 4d). This TR overlaps a multi-allelic repeat variant rs111973244 in dbSNP. When decomposed into multiple biallelic comparisons using conventional GWAS pipelines, the strongest association among rs111973244 alleles only achieves P = 5.49 × 10^-4^ (allele: dup(ATT)_5_, β = 0.11). In contrast, representing the locus as a single TR length variable captures the full repeat-length spectrum and produces a substantially stronger association signal. Conditional analysis by adjusting for the association effect of this TR largely abolished the regional association signals (Fig. S18).

**Figure 4.**
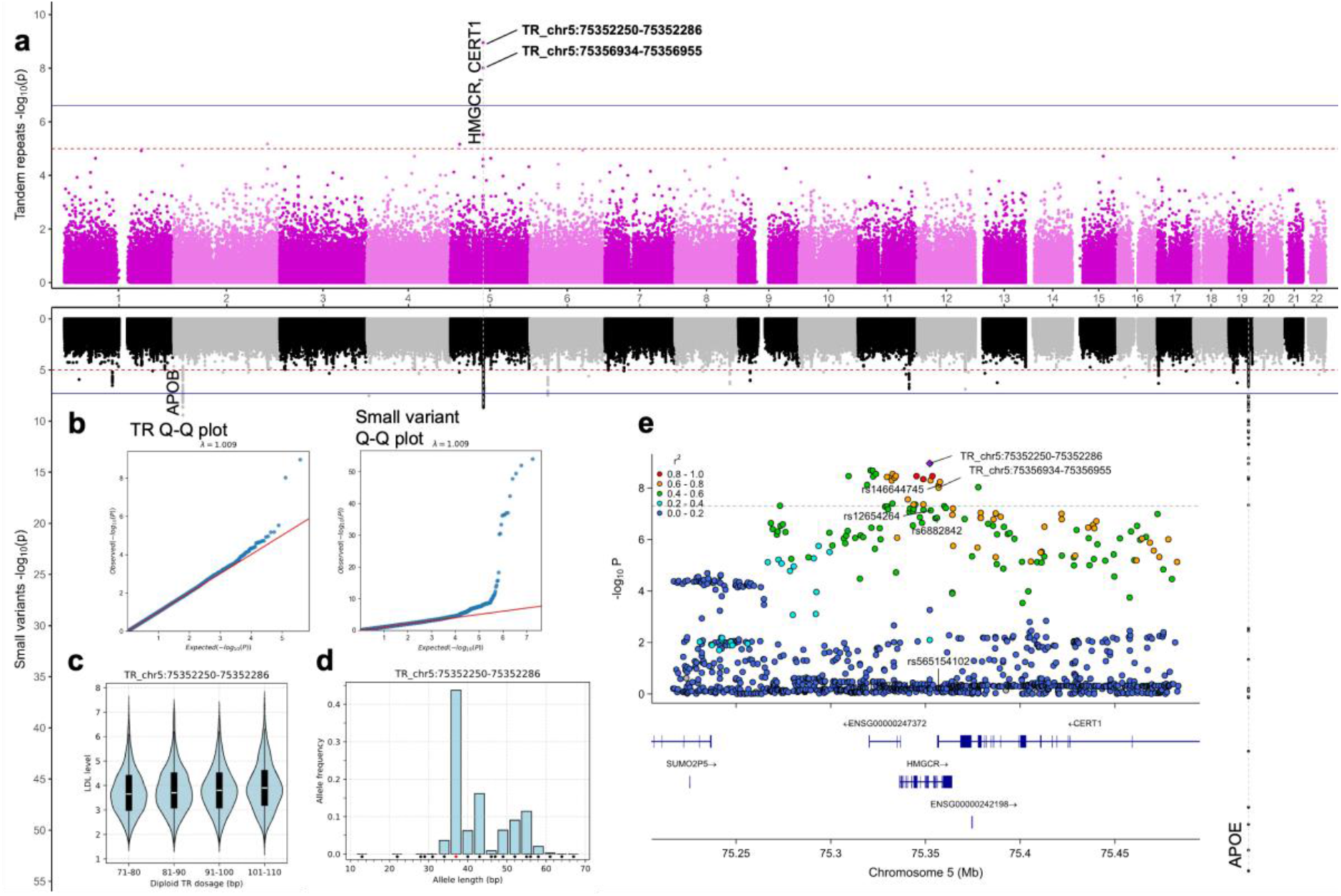
TR-based GWAS identifies a repeat-length association at the HMGCR locus for LDL levels. **a** Mirrored Manhattan plot of TR GWAS (top) and small variant GWAS (bottom) for LDL. **b** Q-Q plots of TR and small variant GWAS for LDL. **c** Association of TR_chr5:75352250-75352286 with LDL levels. **d** Allele length distribution of TR_chr5:75352250-75352286. **e** LocusZoom plot of the HMGCR locus highlighting TR and nearby variants.

A nearby TR variant, TR_chr5:75356934-75356955, located within *HMGCR* intron and the coding C-terminus of *CERT1*, also showed genome-wide association with LDL (P = 9.70 × 10^-9^, β = -0.0069; Fig. S19). This TR is in high LD (r² = 0.79) with TR_chr5:75352250-75352286, but weaker statistical significance (Fig. 5e), indicating that the two signals are correlated. Inspection of this locus shows that its variation reflects composite indel-driven variation, arising from two discrete sequence changes: a 16 bp deletion corresponding to 4 repeat units (rs146644745, P = 7.91 × 10^-9^, β = 0.11) and a 4 bp insertion (rs565154102, P = 6.25 × 10^-1^, β = -0.028). In this case, the deletion variant shows a stronger association than the aggregated TR representation, suggesting that it may be the primary contributor to the observed LDL signal at this locus. This observation further illustrates that the composite TR encoding does not necessarily increase statistical significance but instead provides an alternative representation of local sequence variation. For the remaining six genome-wide significant associations, their association evidence was all weaker than the nearby GWAS signals.

In addition to genome-wide significant loci, 26 TR associations reached a commonly adopted suggestive significance threshold (P < 1 × 10^-5^) but did not meet genome-wide significance (Supplementary Data 9). These loci were not explored further for detailed interpretation, but they provide a set of candidate loci for investigation in further studies. We also conducted GWAS analyses by using small variants. Applying the commonly used genome-wide significance threshold for small variants (P < 5 × 10^-8^), we identified 701 significant associations that fall into 15 independent genomic loci (excluding one singleton artifact) across the five traits, with no significant associations observed for height. Of these, 14 of the 15 loci replicate known GWAS associations for their corresponding traits (Supplementary Data 10-11). The high consistency between the small-variant signals and known GWAS associations serves as an additional layer of control, demonstrating the integrity of the cohort data and the reliability of the analytical framework, which provides additional support for the robustness of the results from the TR GWAS analyses.

## Discussion

In this study, we generated a catalog of 916,274 autosomal TR loci from 9,490 individuals representing three major ethnic groups in Asia. Comparison with the 1KG_H3A-TR catalog revealed 526,891 alleles unique to the SG10K-TR catalog. Although the larger sample size of SG10K_Health cohort likely contributes to this increase rather than reflecting true population-specific alleles, the expanded allele catalog substantially improves TR representation in Asian populations.

We introduced TRDDS as an integrative measure combining allele length variation and frequency differentiation, while TRDDS represents an initial heuristic, it demonstrates the potential for future, more sophisticated approaches to quantify TR population level divergence. Applying this metric, we observed that TR variation accumulates in a manner consistent with broad demographic history and population divergence times. The distribution of population-level TR diversity across genomic annotations reveals a distinct evolutionary dichotomy. On one hand, TR diversity is selectively constrained, likely under strict purifying selection, in core regulatory sequence regions directly governing gene expression, such as CDS, 5’UTRs, and core promoter elements. In contrast, we observed an enrichment of diversity within transcription factor binding regions characterized by low chromatin accessibility. In these context-dependent regulatory elements, TRs likely experience relaxed or balancing selection, acting as rheostatic ‘tuning knobs’ where repeat polymorphisms subtly modulate regulatory dynamics and expression gradients without disrupting core gene functions.

Pathway enrichment analyses of highly diversified TRs consistently highlighted neuronal processes as dominate targets driving population-level TR diversity. We propose these enrichments are directly tied to the unique biological constraints of the nervous system. Because neurons are post-mitotic cells that do not undergo cellular regeneration, and obtaining functional diversity through somatic mutations carries risks of neurodegeneration, while hyper-polymorphic TR variations provide a compact allelic spectrum for functional plasticity. The candidate TR loci under selection on the Chinese and Indian branches we identified in this study validate this neuro-centric model. However, the evolutionary utility of these TRs expands during more recent population splits, reflected by the TR diversity between Chinese and Malay. While core neurological tuning actively continues, recent divergence also incorporates broad structural and metabolic adaptations. This shift is captured systemically by CM-unique pathway enrichments in broader structural and developmental processes, and at the single-locus level by the Malay-specific divergence of a TR within an intron of *C4orf54*, which may have pleiotropic roles in both lipid and neurochemistry. Together, these findings support a unified evolutionary model proposing that while the vast reservoir of TR diversity is fundamentally maintained to regulate long-term neuroplasticity, highly divergent TRs can also be rapidly co-opted to drive localized structural and metabolic adaptations.

Examination of LD patterns across the autosomes revealed substantially weaker LD(TR, small variant) compared with LD(small variant, small variant). This reduced LD may limit the accuracy of imputing TR variation from nearby small variants, particularly for highly polymorphic loci. Nevertheless, a substantial number of TRs were found to be in strong LD with GWAS-associated variants. In particular, 123 candidate functional TRs overlapping CDS showing high LD with trait-associated variants outside CDS were identified, spanning a range of phenotypes. These TRs in CDS may underlie genetic association signals previously attributed to nearby noncoding small variants.

We explored the contributions of TRs to the risk of complex traits by conducting TR GWAS analyses of six anthropometric and lipid traits and identified ten genome-wide significant associations across five traits. Of these 10 loci, two are novel associations that are independent from all the previous GWAS findings, including the BMI-associated TR (TR_chr1:221335333–221335386) and the height-associated *UNC45B* TR (TR_chr17:35177802–35177914). Both loci reside within putative regulatory regions, with the BMI-associated TR overlapping a candidate proximal enhancer and the height-associated TR located within a candidate distal enhancer, suggesting a potential role of TR variation in modulating regulatory activity. While these observations require further functional validation, they highlight the ability of TR GWAS to identify complex variants that cannot be well captured by SNP-based analyses. In addition, we have also identified a TR variant (TR_chr5:75352250–75352286) at *HMGCR* for LDL as a potential causal variant of the previous GWAS association with stronger association evidence. Furthermore, although not independent from previous GWAS findings, the LDL associated coding TR (TR_chr5:75356934-75356955) with genome-wide significant association evidence serve as good candidate for further functional investigations.

Among the ten TRs showing genome-wide significant associations, we identified two variation patterns. The first type is canonical repeat-length variation, exemplified by the *HMGCR* locus (TR_chr5:75352250-75352286), where alleles differ through incremental expansion or contraction of the repeat motif. In such loci, TR-based genotype captures the multiallelic dosage effect of repeat copy number, whereas the overall evidence for genetic effect may become fragmented, when the locus is decomposed into multiple independent biallelic variants for association testing in conventional SNP-based GWAS. The second type is composite indel-involving variation, exemplified by the *UNC45B* locus (TR_chr17:35177802-35177914), that arises from the occurrences of discrete sequence changes within individual repeats. In these cases, TR-based genotype aggregates the combined effects of nearby variants in the repetitive context, enabling more powerful association testing and thus more significant association evidence. Together, these variation patterns captured by TR GWAS provide a basis for novel discoveries beyond conventional SNP-based approaches.

In summary, these findings demonstrate that TR variation is an important and structured component of genetic diversity among Asian populations, and TR diversity highlights biological pathways not typically emphasized in small variant analyses. Furthermore, TR GWAS and LD analyses show that TRs can refine or complement existing trait-associated loci. Several technical considerations were identified in this study. First, long-spanning TRs remain challenging to genotype using current short-read platforms, leading to underrepresentation of long TR insertions in the SG10K-TR catalog. Second, batch effects arising from different sequencing protocols were detected. However, these differences did not impact the overall population structure, and all downstream analyses were subjected to stratified validation to ensure robustness. As long-read sequencing becomes increasingly accessible, future work will be able to resolve long and complex TRs that were underrepresented in the current study. The SG10K-TR catalog also provides a valuable population-specific reference that can serve as a control resource for upcoming disease cohorts, enabling broader exploration of TR contributions to complex traits and disease risk in Asian populations.

## Method

### SG10K_Health TR cohort

The SG10K Health dataset comprises whole-genome sequencing (WGS) data for 9,770 individuals. GRCh38-aligned CRAM files and associated metadata (version r5.3.2) were obtained from the SG10K_Health consortium. For TR genotyping, we focused on a subset of 9,490 samples (referred to as the SG10K_TR cohort) for which sequencing protocol information were available and aligned with the major protocols used in this study. Specifically, this included 6,045 samples prepared with NEBNext Ultra II kit at an average coverage of 15x (15x_PCR-based), 1,922 using the same kit at 30x coverage (30x_PCR-based), and 1,523 samples using the Illumina TruSeq PCR-Free kit at 15x coverage (15x_PCR-free).

### SG10K TR genotyping

#### Single algorithm TR calling

The TR genotyping framework largely followed the EnsembleTR pipeline described by Ziaei Jam *et al.*^11^ Briefly, TR loci were initially genotyped using three callers, that are ExpansionHunter (EH), GangSTR, and HipSTR, and the resulting calls were harmonized using EnsembleTR.

#### EH

EH v5.0.0 was run on each sample and generated per-sample TR VCF file. The definition file used for EH was obtained from https://github.com/Illumina/RepeatCatalogs/blob/master/hg38/variant_catalog.json, and only autosomal loci (N=145,735) were extracted and being genotyped. Per sample VCFs were sorted by bcftools v1.15.1 and merged by MergeSTR v6.0.1.

#### GangSTR

GangSTR v2.5.0 was run on each sample with non-default parameters –grid-threshold 250. The GangSTR definition file was obtained from https://s3.amazonaws.com/gangstr/hg38/genomewide/hg38_ver17.bed.gz, and only non-homopolymer autosomal TR loci (N=801,429) were genotyped. Per sample VCFs were sorted by bcftools and merged by MergeSTR.

#### HipSTR

The 15x_PCR+, 30x_PCR+ and 15x_PCR-groups of the SG10K_TR cohort were further divided into six, four and two smaller batches, respectively. HipSTR v0.6.2 was run on each batch and generated per-batch VCF files using non-default parameters –max-str-len 150. MergeSTR was used to merge per batch VCF files. For HipSTR, an additional correction step was applied using the python script (https://github.com/gymreklab/1000Genomes-STRs/blob/main/Hipstr_correction.py) to further concatenate HipSTR entries that belong to the same locus. Another python script was run to check the format of the merged results to ensure they are applicable for further steps, and unsupported loci were discarded.

### Filter and EnsembleTR calling

#### Level 1 filter

Filter was applied for each TR result VCF using dumpSTR v6.0.1. The following filter criteria were applied to all three caller results to remove loci with low call rate and loci located in segmental duplication regions: --min-locus-callrate 0.75, and –filter-regions hg38_segdup.sorted.bed.gz –filter-regions-names SEGDUP. Segmental duplication regions of hg38 were obtained from https://hgdownload.cse.ucsc.edu/goldenpath/hg38/database/genomicSuperDups.txt.gz and further processed into a BED file. Additional filters were applied for GangSTR VCF: --gangstr-filterspanbound-only and –gangstr-filter-badCI. TR loci failed to meet the filtering criteria were removed by dumpSTR --drop-filtered option.

#### EnsembleTR

EnsembleTR v1.0.0 was run to harmonize the filtered results of ExpansionHunter, GangSTR and HipSTR using default parameters to obtain the consensus VCF.

#### Level 2 filter

A second layer of filter was applied to the EnsembleTR harmonized VCF to further enhance the quality. An modified version of dumpSTR was used to filter the following criteria: --min- locus-callrate 0.75, --min-locus-hwep 0.0000000001, --ensembleTR-min-call-Q 0.2. Known disease-associated TR loci that failed standard filters were selectively retained due to their clinical relevance (Supplementary Methods).

### Annotation and quality control

#### Annotation

Due to the multiallelic nature of TR loci, a two-tiered annotation strategy comprising locus-level genomic context annotation and allele-level functional annotation was employed. First, TR-spanning regions were annotated with gene and regulatory features using BEDTools. Gene annotations were derived from Ensembl release 113 and assigned using the following priority order: coding sequence (CDS), 5′UTR/3′UTR, intronic, non-coding RNA, and intergenic regions. Regulatory annotations were obtained from candidate cis-regulatory elements (cCREs) in the ENCODE Registry via SCREEN^22,23^ (https://downloads.wenglab.org/Registry-V4/GRCh38-cCREs.bed) and assigned according to the following hierarchy: promoter-like signatures (PLS), proximal enhancer-like signatures (pELS), distal enhancer-like signatures (dELS), chromatin accessibility + H3K4me3 (CA-H3K4me3), chromatin accessibility + CTCF (CA-CTCF), chromatin accessibility + transcription factor binding (CA-TF), chromatin accessibility only (CA), and transcription factor binding only (TF). Second, functional consequences of each alternative TR allele were annotated using Ensembl Variant Effect Predictor (VEP) v112.0 with parameters matching those described in Shi *et al.*^13^

#### Mendelian consistency ratio

Identity-by-descent (IBD) analysis was performed using SNP genotypes to infer first-degree relationships based on PLINK 2.0 kinship coefficients. Pairs with kinship among 0.177 to 0.354 and IBS0 < 0.005 were considered likely parent-offspring, and age and gender information were then used to confirm biologically trios, resulting in the identification of 45 parent-offspring trios within the SG10K-TR cohort. For the Mendelian consistency analysis of each trio, consistency checking was performed for TRs that have both alleles valid in all three members, and at least one allele within three members is not the reference allele^11^.

#### Principal component analysis (PCA)

PCA was performed using the sum of both TR allele lengths per individual as genotype input among samples after removing first degree relationships. The analysis was implemented using the PCA function from the scikit-learn Python library.

#### Comparison with 1KG_H3A TR catalog

The 1KG_H3A TR catalog^11^ was obtained from https://ensemble-tr.s3.us-east-2.amazonaws.com/add-vntrs/ensemble_chr*_filtered.vcf.gz. To ensure fair comparison, only TR loci that have exactly same start and end positions between both catalogs were extracted for allele counting.

### TR diversity among three ethnic groups

#### Population level TR diversity

##### Rst

Rst values were calculated following the definition by Slatkin^21^, representing the proportion of variance between groups over the total variance:

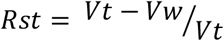

where *Vt* is the total variance, and *Vw* is the weighted within-group variance. The between-group variance is defined as the difference between the total and within-group variances.

##### TRDS

TRDS was computed as the two-way Wasserstein distance between allele length distributions of two populations, following the method described in Cui *et al.*^14^ This was implemented using the scipy.stats.wasserstein_distance function by Python scipy package.

##### TRDDS

The TRDDS metric created by this study is defined as the product of Rst and TRDS. A detailed mathematical rationale and additional properties of TRDDS are provided in Supplementary Note 1. For TRDDS between two groups, it is the direct product of Rst and TRDS between two groups. For multi-group TRDDS, for example, the overall TRDDS level among Chinese, Malay and Indian, it is calculated as the product of multi-group Rst and sum of the pairwise TRDS values:

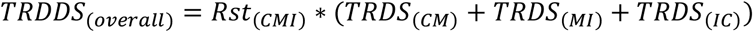

##### Enrichment analysis

For each population pair (CM, MI, and CI), genes overlapping the top 0.5% of TR loci ranked by TRDDS were extracted for Gene Ontology (GO) enrichment analysis. Non-coding RNA (ncRNA) genes were excluded. GO enrichment was performed using the R package clusterProfiler, with the pvalueCutoff set to 0.01.

#### TR loci under selection

##### TR-PBS

The Population Branch Statistic (PBS) framework was adapted for TR based on pairwise TRDDS values to estimate the magnitude of genetic divergence occurring specifically along a single population lineage relative to a shared ancestral node. For each pTR, the TR-PBS for a specific lineage, e.g., the Chinese branch PBS(C), was calculated using the pairwise TRDDS values as follows:

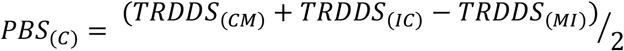

The PBS(M) and PBS(I) values were calculated symmetrically for their respective branches. For each pTR, the branch corresponding to the max TR-PBS value (PBS_max_) among the three PBS values designated the dominant evolutionary branch. Candidate TRs under selection were defined as the 0.1% of loci based on PBS_max_, subjected to batch validation (Supplementary Methods).

##### Fold Change

To filter out ambiguous topologies or shared ancestral divergence events, a fold change (FC) measure was applied, defined as the ratio of PBS_max_ to the second-highest PBS value for each pTR. A threshold of FC ≥ 2 was applied to the candidate TRs. Finally, the relative strength of divergence for retained TR candidates was assessed using an Impact Score, calculated as *PBS_max_* × *log*_2_(*FC*).

#### Linkage disequilibrium

##### Relative diploid TR dosage

TR genotype was recoded as relative diploid TR dosage (shortened as TR dosage hereafter) for LD calculation. For each TR locus with reference repeat length r, the TR dosage was calculated as the sum of allele-length differences from the reference:

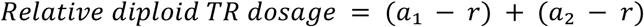

where a₁ and a₂ are the two TR allele lengths carried by an individual at a TR locus.

For example, at a TR locus with a 9 bp reference length, a genotype of 12 bp / 15 bp yields a diploid dosage of (12 − 9) + (15 − 9) = 9.

##### Length-based LD calculation

TR genotypes of each individual were recoded as the TR dosage. For each TR locus, the TR dosage category with frequency < 0.01 were excluded from LD calculation, and if a TR locus does not have more than one TR dosage category with frequency ≥ 0.01, the whole TR locus was discarded from pairwise LD calculation. Small variant genotypes were encoded as 0, 1, or 2, representing the number of alternative alleles. LD was computed as the squared Pearson correlation coefficient (r²) using the .corr() function in Python pandas library.

##### TR tagged by GWAS variants

GWAS catalog v1.0.2 was downloaded from https://www.ebi.ac.uk/gwas/api/search/downloads/alternative. Variants with valid rsIDs (extracted from the “SNPS” column) were intersected with TR-SNP LD pairs. TRs were considered tagged by a GWAS variant if the corresponding r² ≥ 0.6. For population significant TR - GWAS variant pairs, we additionally require the r² < 0.3 for the population(s) that are not significant, and LD was further checked using non-filtered TR genotypes and genotype frequency of 0.05 to ensure the significance of LD is robust to genotype frequency.

#### TR GWAS

##### Sample filtering

Sample filtering involved three key steps. First, the GUSTO cohort which contains infants within the SG10K-TR cohort was excluded. Second, individuals with first-degree relationships were removed. Third, samples failing quality thresholds based on small variant genotypes were excluded: sample call rate < 0.95, heterozygosity rate < mean + 3std. After sample filters, the GWAS cohort contains 6,458 individuals.

##### Phenotype normalization

Essential normalization was applied for the six continuous phenotypes-body mass index (BMI), height, high-density lipoprotein (HDL), low-density lipoprotein (LDL), total cholesterol (TC), and triglycerides (TG). The TG values were log-transformed followed by rank-based inverse normal transformation. The remaining five phenotypes had log transformation only. After transformation, z-score normalization was applied for each trait, and outliers with z-score > 3 were discarded.

##### Small variant GWAS

Small variants meeting quality criteria were included for association testing: locus call rate > 0.95, Hardy-Weinberg equilibrium p-value > 1×10⁻^10^, and MAF > 0.01. Association analyses for all six phenotypes were performed using PLINK2’s generalized linear model (--glm) with covariates including sex, age, and small variant based PC1-PC5.

##### TR GWAS

For each TR locus, consistent with the LD workflow, TR genotypes were recoded as TR dosage. This encoding models the additive effect of total repeat length on the phenotype. Additional TR-specific filters were applied beyond the initial SG10K-TR catalog processing for TR GWAS. Because association testing was performed on TR dosage rather than individual alleles, frequency filtering was applied at the dosage category level. Specifically, TR loci were excluded if no non-reference dosage category had a frequency ≥ 1% in the study population. For retained loci, dosage categories with frequency < 1% were excluded from regression analysis to avoid instability due to sparse observations. Furthermore, TR loci were required to exhibit at least one repeat unit difference among dosage categories with frequency ≥ 1% to ensure meaningful length variation for association testing. Only loci satisfying these criteria were carried forward for regression analysis. Association testing was performed using ordinary least squares regression (Python statsmodels sm.OLS()), adjusting for sex, age, and the first five principal components derived from TR genotypes (TR PC1-PC5).

##### LocusZoom

LocusZoom plots of top TR signals were generated using the R package locuszoomr^49^. Gene regions were annotated based on Homo Sapiens Ensembl release 110. The LD values between the top TR signals and nearby small variants or TRs were calculated following the approach described above.

## Acknowledgement

This study made use of data generated as part of the Singapore National Precision Medicine program funded by the Industry Alignment Fund (Pre-Positioning) (IAF-PP:H17/01/a0/007). The computational work for this article was partially performed on resources of the National Super Computing Centre, Singapore (https://www.nscc.sg). This study made use of data / samples collected in the following cohorts in Singapore: (1) The Health for Life in Singapore (HELIOS) study at the Lee Kong Chian School of Medicine, Nanyang Technological University, Singapore (supported by grants from a Strategic Initiative at Lee Kong Chian School of Medicine, the Singapore Ministry of Health under its Singapore Translational Research Investigator Award (NMRC/STaR/0028/2017) and the IAF-PP: H18/01/a0/016; (2) The Growing up in Singapore Towards Healthy Outcomes (GUSTO) study, which is jointly hosted by the National University Hospital (NUH), KK Women’s and Children’s Hospital (KKH), the National University of Singapore (NUS) and the Singapore Institute for Clinical Sciences(SICS), Agency for Science Technology and Research (A*STAR) (supported by the Singapore National Research Foundation under its Translational and Clinical Research(TCR) Flagship Programme and administered by the Singapore Ministry of Health’s National Medical Research Council (NMRC), Singapore - NMRC/TCR/004-NUS/2008; (3)The Singapore Epidemiology of Eye Diseases (SEED) cohort at Singapore Eye Research Institute (SERI) (supported by NMRC/CIRG/1417/2015; NMRC/CIRG/1488/2018;NMRC/OFLCG/004/2018); (4) The Multi-Ethnic Cohort (MEC) cohort (supported by NMRC grant 0838/2004; BMRC grant 03/1/27/18/216; 05/1/21/19/425; 11/1/21/19/678, Ministry of Health, Singapore, National University of Singapore and National University HealthSystem, Singapore); (5) The SingHealth Duke-NUS Institute of Precision Medicine(PRISM) cohort (supported by NMRC/CG/M006/2017_NHCS; NMRC/STaR/0011/2012,NMRC/STaR/0026/2015, Lee Foundation and Tanoto Foundation); (6) The TTSH Personalised Medicine Normal Controls (TTSH) cohort funded (supported by NMRC/CG12AUG17 and CGAug16M012).

The views expressed are those of the author(s) are not necessarily those of the National Precision Medicine investigators, or institutional partners. We thank all investigators, staff members and study participants who made the National Precision Medicine Project possible.

## Data availability

The SG10K_Health dataset used in this study was obtained under Data Access Application NPM00035 through the National Precision Medicine (NPM) program Phase I Data Access Committee (DAC). Due to participant consent restrictions and data privacy regulations, the dataset is available under controlled access. Interested researchers seeking access to the dataset are required to submit a data access request for evaluation by the NPM Phase I DAC. Further information can be found at https://www.a-star.edu.sg/gis/our-science/precision-medicine-and-population-genomics/npm/data-access.

## Code availability

Scripts used for data processing and analysis in this study are available in the GitHub repository (https://github.com/c-BIG/SG10K-TR_manuscript).

## Ethics

This project is approved by the NPM Data Access Committee (DAC) with project ID: NPM00035.

## Author contributions

J.Liu, and N.B. conceived and supervised the project. Q.J. and L.W conducted upstream TR calling. Q.J. conducted downstream data analysis. Q.J., and M.L., contributed to manuscript writeup with contributions from all authors. L.W., F.Z., and H.T., P.S., and Z.L. contributed to data analysis and interpretation. E.W. contributed to project management. P.T. contributed to study supervision and manuscript review. P.T., X.S, J.N., J.Lee, C.Y.C., M.L.C., W.K.L., C.W.L.C., N.K., Y.S.C., W.C.S., C.W.L., and the SG10K_Health Consortium contributed to data collection and production.

## SG10K_Health Consortium

Tin Aung^1,2^, Claire Bellis^3,4^, Nicolas Bertin^5^, Jin Fang Chai^6^, John C Chambers^7,8,9^, Miao Ling Chee^1^, Ching-Yu Cheng^1,2,10,11^, Paul Chung Pui Cheng^12^, Wen Jie Chew^13^, Calvin Woon Loong Chin^14,15^, Yap Seng Chong^16,17^, Stuart Alexander Cook^18,19,20^, Paul Eillot^8,21^, Johan G Eriksson^22,23,24,25^, Peter D Gluckman^17,26^, Liuh Ling Goh^27^, Maxime Hebrard^5^, Pritesh Rajesh Jain^7^, Saumya S Jamuar^28,29,30,31^, Justin Jeyakani^5^, Rodrigo Toro Jimenez^5^, Neerja Karnani^22,32,33^, Tat Hung Koh^3^, Eng Sing Lee^21,34^, Jimmy Lee^21,35^, Yung Seng Lee^16,36^, Khai Pang Leong^27^, Hengtong Li^1,11^, Zhihui Li^5^, Chia Wei Lim^27^, Tock Han Lim^21,37^, Weng Khong Lim^28,38,39^, Bitong Clarabelle Alexandrine Lin^3^, Jianjun Liu^40,41^, Marie Loh^7,8,42^, Dorrain Low^7^, Roberto-Tirado Magalanes^5^, Sebastian Maurer-Stroh^43,44^, Theresia Mina^7^, Shiqi Mok^12^, Hong Kiat Ng^7^, Joanne Ngeow^7,45,46^, Jack Ling Ow^5^, Qingsheng Peng^1,2^, Shyam Prabhakar^47^, Chee Jian Pua^19^, Elio Riboli^8,21^, Charumathi Sabanayagam^1,2^, Nilanjana Sadhu^7^, Yee Yen Sia^40^, Wey Ching Sim^27^, Xueling Sim^6^, E Shyong Tai^6,9,41^, Joanna Hui Juan Tan^5^, Patrick Tan^9,48,49^, Erwin Tantoso^50^, Darwin Tay^7^, Yik Ying Teo^6^, Yih Chung Tham^1,10,11^, Li-xian Grace Toh^27^, Pi Kuang Tsai^27^, Rob M van Dam^6,51^, Chandra Verma^52,53,54^, Xiaoyan Wang^7^, Tien Yin Wong^1,2^, Can Can Xue^1^, Chengxi Yang^19^, Fabian Yap^49,55^, Khung Keong Yeo^14,28,49^

Authors are ordered alphabetically by last name.

^1^Singapore Eye Research Institute, Singapore National Eye Centre, Singapore, ^2^Ophthalmology & Visual Sciences Academic Clinical Program, Duke-NUS Medical School, Singapore, ^3^Extract Platform, Genome Institute of Singapore, Agency for Science, Technology and Research (A*STAR) Singapore, Singapore, ^4^Centre for Genomics and Personalised Health, Genomics Research Centre, School of Biomedical Sciences, Queensland University of Technology, Brisbane, Australia, ^5^NPM-Genomic Intelligence & Informatics Engine (NPM-GINIE), Genome Institute of Singapore, Agency for Science, Technology and Research (A*STAR) Singapore, Singapore, ^6^Saw Swee Hock School of Public Health, National University of Singapore and National University Health System, Singapore, ^7^Population and Global Health, Lee Kong Chian School of Medicine, Nanyang Technological University, Singapore, ^8^Department of Epidemiology and Biostatistics, Imperial College London, London, UK, ^9^Precision Health Research, Singapore (PRECISE), Singapore, ^10^Department of Ophthalmology, Yong Loo Lin School of Medicine, National University of Singapore and National University Health System, Singapore, ^11^Centre for Innovation and Precision Eye Health, Yong Loo Lin School of Medicine, National University of Singapore and National University Health System, Singapore, ^12^Laboratory of Complex Disease Genetics, Genome Institute of Singapore, Agency for Science, Technology and Research (A*STAR) Singapore, Singapore, ^13^Clinical Research & Innovation Office, Tan Tock Seng Hospital, Singapore, ^14^Department of Cardiology, National Heart Centre Singapore, Singapore, ^15^Cardiovascular Academic Clinical Program, Duke-NUS Medical School, Singapore, ^16^Department of Obstetrics & Gynaecology, Yong Loo Lin School of Medicine, National University of Singapore and National University Health System, Singapore, ^17^Institute for Human Development and Potential, Agency for Science, Technology and Research (A*STAR) Singapore, Singapore, ^18^Cardiovascular and Metabolic Disorders Program, Duke-NUS Medical School, Singapore, ^19^National Heart Research Institute Singapore, National Heart Centre Singapore, Singapore, ^20^MRC Laboratory of Medical Sciences, London, UK, ^21^Lee Kong Chian School of Medicine, Nanyang Technological University, Singapore, ^22^Human Development, Institute for Human Development and Potential, Agency for Science, Technology and Research (A*STAR) Singapore, Singapore, ^23^Department of Obstetrics & Gynaecology, Yong Loo Lin School of Medicine, National University of Singapore and National University Health System, ^24^Department of General Practice and Primary Health Care, Folkhalsan Research Center, Finland, ^25^Department of General Practice and Primary Health Care, University of Helsinki, Finland, ^26^Liggins Institute, University of Auckland, New Zealand, ^27^Centre for Precision and Genomic Medicine, Tan Tock Seng Hospital, Singapore, ^28^SingHealth Duke-NUS Institute of Precision Medicine, Singapore, ^29^SingHealth Duke-NUS Genomic Medicine Centre, Singapore, ^30^Genetics Service, Department of Paediatrics, KK Women’s and Children’s Hospital, Singapore, ^31^Paediatric Academic Clinical Program, Duke-NUS Medical School, Singapore, ^32^Clinical Data Engagement, Bioinformatics Institute, Agency for Science, Technology and Research (A*STAR) Singapore, Singapore, ^33^Department of Biochemistry, National University of Singapore, Singapore, ^34^Clinical Research Unit, National Healthcare Group Polyclinics, Singapore, ^35^Research Division, Institute of Mental Health, Singapore, ^36^Khoo Teck Puat-National University Children’s Medical Institute, National University Hospital, Singapore, ^37^National Healthcare Group Eye Institute, Tan Tock Seng Hospital, Singapore, ^38^Cancer & Stem Cell Biology Program, Duke-NUS Medical School, Singapore, ^39^Laboratory of Genome Variation Analytics, Genome Institute of Singapore, Agency for Science, Technology and Research (A*STAR) Singapore, Singapore, ^40^Laboratory of Human Genomics, Genome Institute of Singapore, Agency for Science, Technology and Research (A*STAR) Singapore, Singapore, ^41^Department of Medicine, Yong Loo Lin School of Medicine, National University of Singapore and National University Health System, Singapore, ^42^Laboratory of DNA methylation and Population Epigenetics, Genome Institute of Singapore, Agency for Science, Technology and Research (A*STAR) Singapore, Singapore, ^43^Protein Sequence Analysis, Bioinformatics Institute, Agency for Science, Technology and Research (A*STAR) Singapore, Singapore, ^44^Yong Loo Lin School of Medicine, National University of Singapore and National University Health System, Singapore, ^45^Cancer Genetics Service, Division of Medical Oncology, National Cancer Centre, Singapore, ^46^Oncology Academic Clinical Program, Duke-NUS Medical School, Singapore, ^47^Laboratory of Systems Biology and Data Analytics, Genome Institute of Singapore, Agency for Science, Technology and Research (A*STAR) Singapore, Singapore, ^48^Genome Institute of Singapore, Agency for Science, Technology and Research (A*STAR) Singapore, Singapore, ^49^Duke-NUS Medical School, Singapore, ^50^Data Management, Bioinformatics Institute, Agency for Science, Technology and Research (A*STAR) Singapore, Singapore, ^51^Departments of Exercise and Nutrition Sciences and Epidemiology, Milken Institute School of Public Health, The George Washington University, DC, USA, ^52^Atomistic Simulations and Design in Biology, Bioinformatics Institute, Agency for Science, Technology and Research (A*STAR) Singapore, Singapore, ^53^School of Biological Sciences, Nanyang Technological University, Singapore, Singapore, ^54^Department of Biological Sciences, National University of Singapore, Singapore, ^55^Department of Paediatrics, KK Women’s and Children’s Hospital, Singapore

